# Utilize State Transition Matrix Model to Predict the Novel Corona Virus Infection Peak and Patient Distribution

**DOI:** 10.1101/2020.02.16.20023614

**Authors:** Ke Wu, Junhua Zheng, Jian Chen

**Author notes:** Corresponding: Jian Chen, CreditWise Technology Co., Ltd. Floor 4-5, Section B, Building 1 Tianfu 5^th^ Ave., Chengdu Hi-tech Zone Chengdu, 610041, China., Junhua Zheng, Department of Evidence-based medicine, Shanghai General Hospital, Shanghai Jiao, Tong University, School of Medicine, Haining Road 100, 200000, China.

## Abstract

**Background:** Since December 2019, a pneumonia caused by the 2019 novel coronavirus (2019-nCoV) has broken out in Wuhan, Hubei province, China. The continuous rising of infected cases has imposed overwhelming pressure on public health decision and medical resource allocation in China. We managed to forecast the infection peak time in Hubei province and the severe and critical case distribution.

**Methods:** We used data resource according to cases reported by the National Health Commission of the People’s Republic of China (Jan 25, 2019, to Feb 28, 2020) as the training set to deduce the arrival of the peak infection time and the number of severe and critical cases in Wuhan on subsequent days. Medical observation, discharge, infected, non-Severe, infected and severe, cure and death data were collected and analyzed. Using this state transition matrix model, we will be able predict when the inflection peak time (the maximum open infection cases) in Hubei Province will occur. Also, we can use this model to predict the patient distribution (severe, non-severe) to better allocate medical resource. Under relative pessimistic scenario, the inflection peak time is April 6-April 14. The numbers of critically ill and critically ill patients will lie between 8300-9800 and 2200-2700, respectively.

**Results:** In very optimistic scenarios (daily NCC decay rate of −10%), the peak time of open inflection cases will arrive around February 23-February 26. At the same time, there will be a peak in the numbers of severely ill and critically ill patients, between 6800-7200 and 1800-2000, respectively. In a relative optimistic scenario (daily NCC decay rate of −5%), the inflection case peak time will arrive around February 28-March 2. The numbers of critically ill and critically ill patients will lie between 7100-7800 and 1900-2200, respectively. In a relatively pessimistic scenario (daily NCC decay rate of −1%), the inflection peak time does not arrive around the end of March. Estimated time is April 6-April 14. The numbers of critically ill and critically ill patients will lie between 8300-9800 and 2200-2700, respectively. We are using the diagnosis rate, mortality rate, cure rate as the 2/8 data. There should be room for improvement, if these metrics continue to improve. In that case, the peak time will arrive earlier than our estimation. Also, the severe and critical case ratios are likely to decline as the virus becomes less toxic and medical conditions improve. If that happens, the peak numbers will be lower than predicted above.

**Conclusion:** We can infer that we are still not close to the end of this outbreak and the number of critically ill patients is still climbing. Assisting critical care resources in Hubei province requires the government to consider further tilt, and it is vital to make reasonable management of doctors and medical assistance systems to curb the transmission trend.

## Introduction

Novel coronavirus pneumonia (NCP) caused by zoonotic 2019 novel coronavirus (2019-nCoV) outbreaks in China during early December 2019^1^.The Chinese Government has progressively implemented several measures to stop the spread of the epidemic^2^. Given the high prevalence and wide distribution of coronaviruses, the condition will continue for some time^3^. As of February 10th, 2020, 37,626 laboratory-confirmed cases and 1016 death cases have been documented in China. Pragmatic effectiveness trials are increasingly recognized as an essential component of medical evidence and good prediction models can help formulate scientific prevention and treatment programs^4, 5^.

In the recent past, two other novel coronaviruses (CoVs), severe acute respiratory syndrome coronavirus (SARS-CoV; in 2002) and Middle East respiratory syndrome coronavirus (MERS-CoV; in 2012) have emerged, and the world is placed on high alert^6, 7^. They have similar symptoms, while SARS-CoV and MERS-CoV have low potential for sustained community transmission^8^. Infections and deaths of SARS-CoV and MERS-CoV are less than 2019-nCoV, thus the previous prediction model is no longer suitable for 2019-nCoV^9^.

In this study, we provide State Transition Matrix Model to predict the 2019-nCoV infection peak and patient distribution. More importantly, from a public health viewpoint, we then estimate the risk metrics (infectivity, severity and lethality) of the NCP. We set up six different scenarios, in order to control for model error. So, we can identify the close contact/patient’s state by utilizing a state vector at any moment.

## Methods

### Study Population

Medical observation, discharge, infected, non-Severe, infected and severe, cure and death data and corresponding information released by the National Health Commission of the People’s Republic of China from Jan 25, 2019, to Feb 28, 2020 were analyzed.

### State Transition Matrix Model

State transition matrix modeling is a well-regarded approach widely applied in clinical decision analysis based on computer simulation. For estimating the infection peak time and the scale of severe and critical cases in subsequent days, we chose the Markov model cohort simulation.

### Parameter Selection and Estimate

In order to estimate the risk metrics (infectivity, severity, lethality) of the NCP, we build a state transition matrix model as the following.

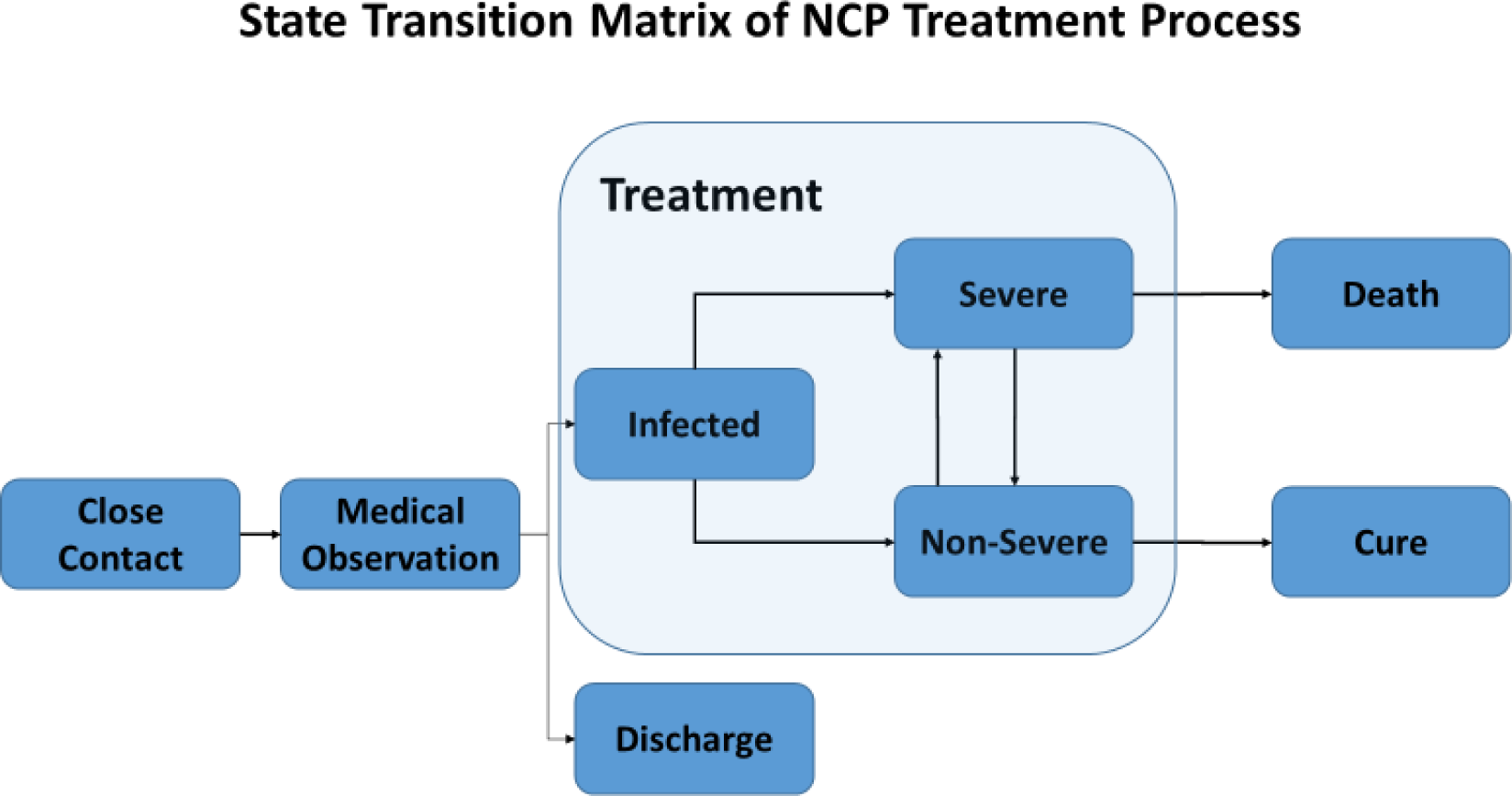

We define the states in this model. Medical Observation (MO) is a close contact of a known infected patient is identified and put into medical observation. In the next day, outcome could be any of the three: confirmed infection, discharged without infection, or stay in MO. Discharge (dis) is a terminal state for a close contact, until he or she becomes another incident of close contact again. Infected is an intermediate state, where the patient becomes a confirmed infected case. The outcome is binary: severe, or non-severe. And the outcome is revealed immediately. Non-Severe (INS) is the patient also has three possible outcomes in the next day: cure, severe, or stay in non-severe. Infected and Severe (IS), the patient has three possible outcomes in the next day: death, non-severe, or stay in severe. Cure (Cu) is also a terminal state for the patient. Death (D) is a terminal state for the patient. So, at any moment, we can identify the close contact/patient’s state by utilizing a state vector, defined as the following:

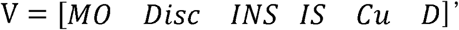

Where each element of the vector stands for one state in the same sequentially arranged order as mentioned above. Please note that infected itself is not an independent state, since the outcome is revealed instantaneously, so we combine Infected with Severe and Non-Severe.

For each person, the state vector can only have one element with value of 1, and the other elements all have value of zero. For example, if a patient is currently in state “infected and severe”, the state vector for her is [0 0 1 0 0 0]’. The next day, her state vector could become either [0 0 0 0 1 0]’ (cure) or stay the same.

For the sample population, the state vector is defined as the count of people in each state. For example, if there are 100 patients being treated today, out of which 10 are severe, and 90 are non-severe. The state vector for this sample population is [0 0 90 10 0 0]’.

Let’s define the state transition matrix as the following:

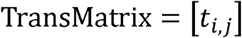

Where

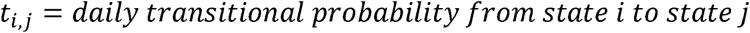

Suppose we have a state vector V(t) for a sample population at time t, how do we predict the state vector V(t+1) in the next day?

Apply simple linear algebra, we can get the following equation:

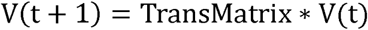

Since the head count of a certain state comes from itself, all other possible transitions into the state (e.g. INS has two possible income states, MO and IS), minus the outcome states (IS, and Cu).

If we want to predict for N period, the equation becomes the following:

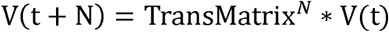

If the population is limited and the transition matrix is stationary, the above formula will be sufficient in predicting all future outcomes. In our case, the population is not fixed, so we need to introduce the additional input into the population: new close contacts.

Every day, new close contacts are added to the medical observation pool, as people already in the pool will gradually be discharged or confirmed of infection.

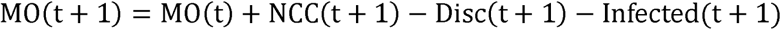

Also we assume NCC will gradually decay as quarantine measures are put into effect.

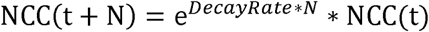

Using this state transition matrix model, we will be able predict when the inflection peak time (the maximum open infection cases) in Hubei Province will occur. Also, we can use this model to predict the patient distribution (severe, non-severe) to better allocate medical resource.

Although there is an intermediate state during the above hospitalization: non-severe illness (the new standard is broken down into mild and normal), severe illness (which can also be divided into general critical and critical), due to the lack of intermediate state transfer probability, we combine the entire hospital period into a therapeutic state, in order to keep the model simple. This minimizes the need for only the following five parameters.

- Decay Rate of New Close Contacts (NCC), defined as ln(NCC(t)/NCC(t-1));
- Discharge Rate from Medical Observation (MO), defined as Discharged(t)/MO(t-1)
- Transitional Probability of Medical Observation −> Confirmed Infection, defined as New Infection Cases (t)/MO(t-1)
- Transitional Probability of Treatment −> Death, defined as New Death Incidents(t) / Treatment(t-1)
- Transitional Probability of Treatment −> Cure, defined as New Cure Incidents (t) / Treatment(t-1)

In order to estimate the count of open severe cases and critical cases, we need two more parameters:

- Ratio of Severe Cases
- Ratio of Critical Cases

We collected all available data from Caixin Data (a subsidiary of Caixin Group), who collects the original data from National Health Commission. We collected supplementary data from Hubei Health Commission. The data period starts from 2019/12/31, and ends on 2020/2/11. Data is updated on a daily basis.

Next, we describe how we estimate these parameters from empirical probabilities.

### Scenario Setup

We set up six different scenarios, in order to control for model error. Two of the scenarios are very optimistic, two of them are relatively optimistic, and two of them are relatively pessimistickan (Table 1).

## Results

### Parameter Estimation

This decay rate is closely linked to R0 and is directly estimated from empirical data. The 8-day average is −6%, but it is volatile, so we will test −1%, −5%, −10% respectively (Figure 1A). Since we could not obtain the one-day probability of lifting medical observation in Hubei Province, we used national rate for reference. The latest one-day probability is 17% and the 10-day moving weighted average is 13%. Considering that the probability of Hubei Province may be lower than that of the whole country, we test it with 17% and 13%, respectively. (Figure 1B). In Hubei Province, the latest one-day probability of transition from medical observation to confirmed infection is 2.15%, with a weighted average of 3.94% over the last 10 days, ending at 2/11. As this probability continues to decline, we use the average (3.04%) of these two values as our model parameter to be cautiously optimistic (Figure 1C). The latest single-day mortality rate was 0.33% and the 10-day moving weighted average was 0.41%. As this probability continues to decline, we use the average (0.37%) of these two values as our model parameter to be cautiously optimistic (Figure 1D). The latest one-day cure rate is 1.46% and the 10-day moving weighted average is 1.27%. As this probability continues to decline, we use the average (1.37%) of these two values as our model parameter to be cautiously optimistic (Figure 1E). From the historical data, the proportion of critical cases is relatively stable; the proportion of severe cases fluctuates more. As these two probabilities continues to fluctuate, we also use the average (17.56%, and 4.82%) of the latest value and 10-day moving weighted average as our model parameters to put more weight on recent observation. (Figure 1F). The parameters we use in our forecast are listed in Table 2.

**Figure 1.**
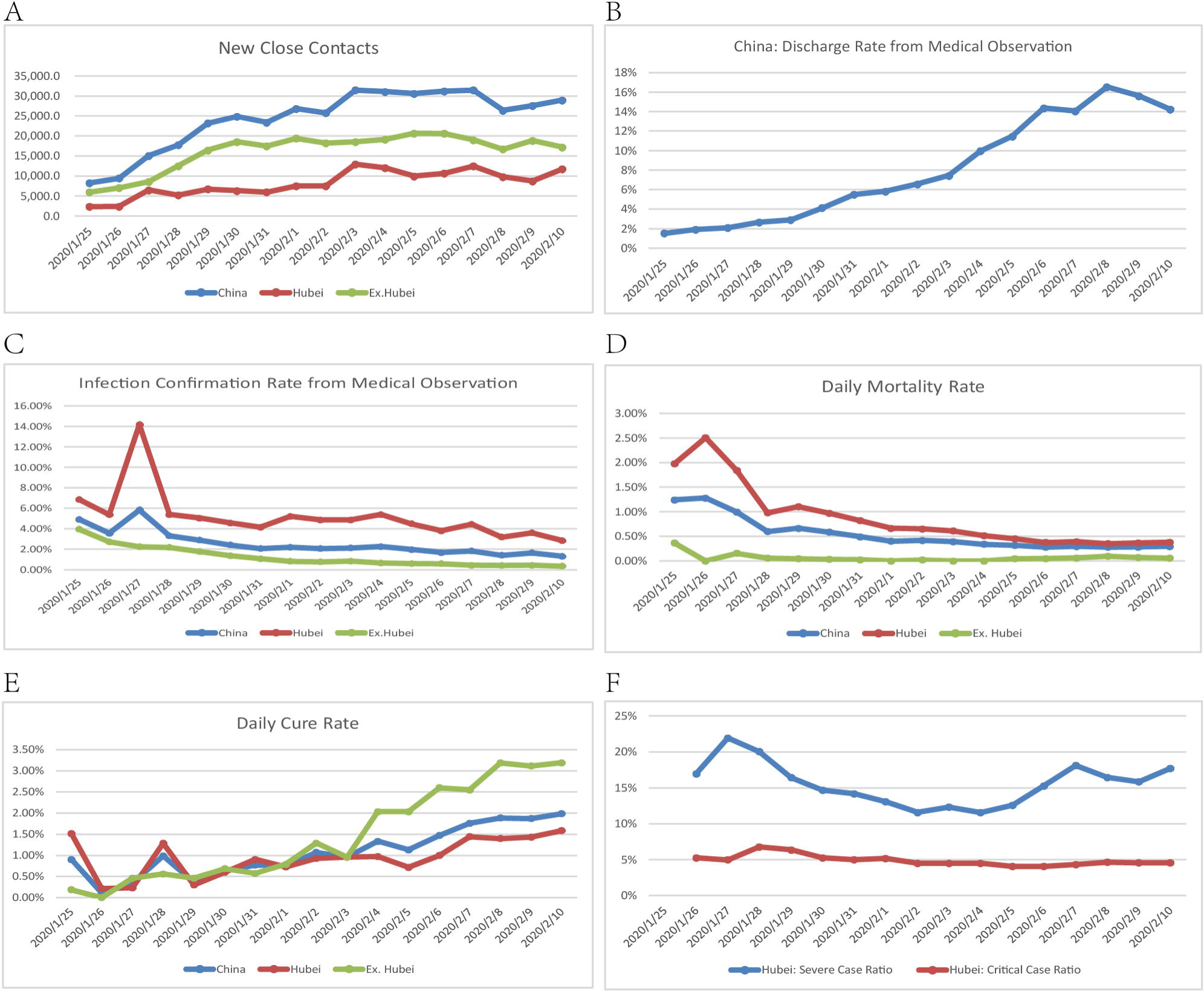
Parameter Estimation of forecast model. A. The trend of new close contacts among Hubei, outside Hubei and the total of China. B. 3 Dynamic profile of discharge rate from medical observation in China. C. The infection confirmation rate from medical observation to in Hubei, outside Hubei and the total of China. D. The daily mortality rate of NCP among Hubei, outside Hubei and the total of China. E. The daily cure rate of NCP among Hubei, outside Hubei and the total of China. F. The daily proportion of severe and critical NCP in Hubei.

### Peak Time Estimation

In very optimistic scenarios (daily NCC decay rate of −10%), the peak time of open inflection cases will arrive around February 23-February 26. At the same time, there will be a peak in the numbers of severely ill and critically ill patients, between 6800-7200 and 1800-2000, respectively (Figure 2A and 2B). In a relative optimistic scenario (daily NCC decay rate of −5%), the inflection case peak time will arrive around February 28-March 2. The numbers of critically ill and critically ill patients will lie between 7100-7800 and 1900-2200, respectively (Figure 2C and 2D). In a relatively pessimistic scenario (daily NCC decay rate of −1%), the inflection peak time does not arrive around the end of March. Estimated time is March 25 − April 2. The numbers of critically ill and critically ill patients will lie between 8300-9800 and 2200-2700, respectively (Figure 2E and 2F). We are using parameters with 50% weight on the diagnosis rate, mortality rate, cure rate as of 2/11. There should be room for improvement, if these metrics continue to improve. In that case, the peak time will arrive earlier than our estimation. Also, the severe and critical case ratios are likely to decline as the virus becomes less toxic and medical conditions improve. If that happens, the peak numbers will be lower than predicted above (Figure 2G and2H). The parameters we use in our forecast are listed in Table 2.

**Figure 2.**
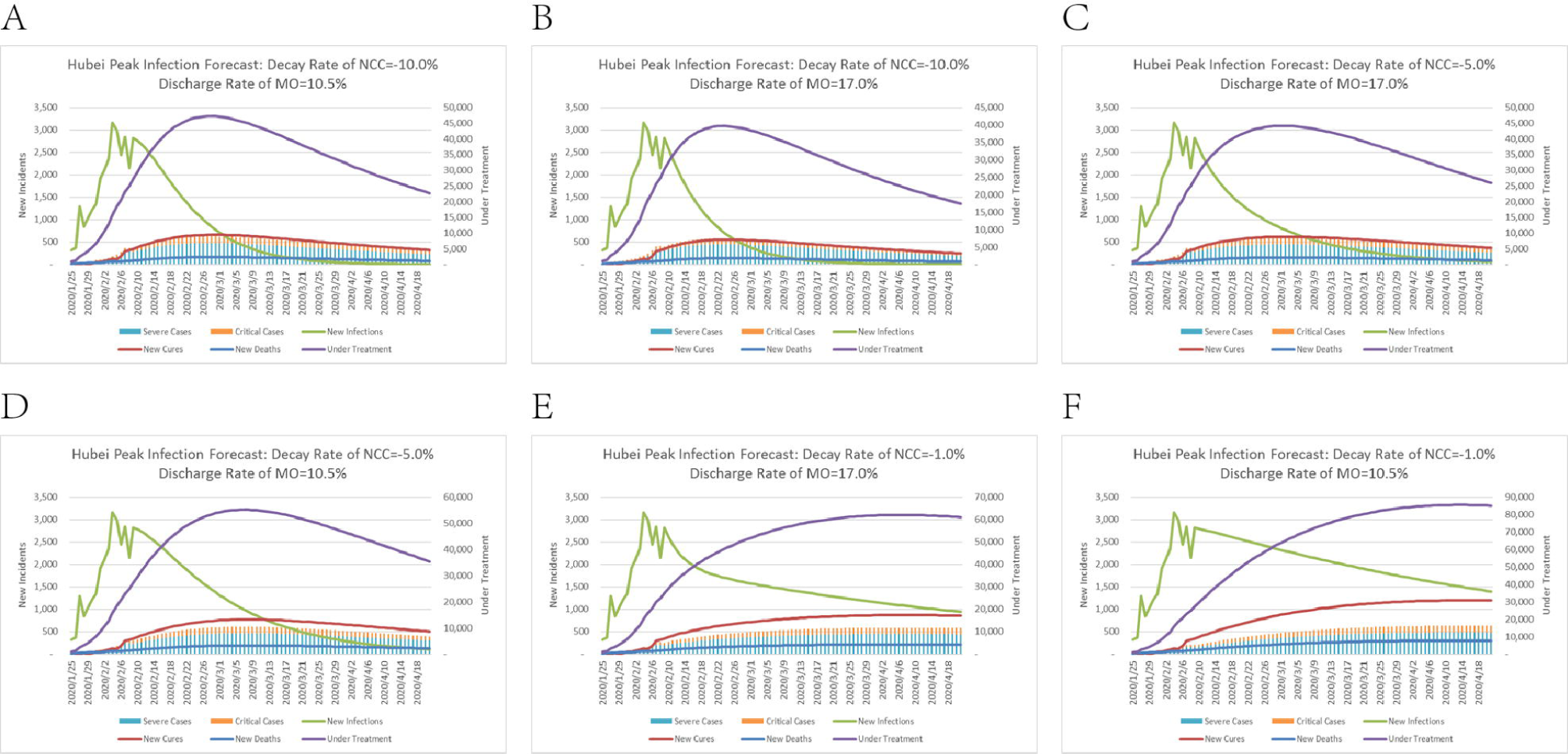
Peak Time Estimation. A. The peak infection time and patient distribution forecast of Hubei (Parameter Set: Decay Rate of NCC = −10%, Discharge Rate of MO = 10.5%). B. The peak infection time and patient distribution forecast of Hubei (Parameter Set: Decay Rate of NCC = −10%, Discharge Rate of MO = 17%). C. The peak infection time and patient distribution forecast of Hubei (Parameter Set: Decay Rate of NCC = −5%, Discharge Rate of MO = 17%). D. The peak infection time and patient distribution forecast of Hubei (Parameter Set: Decay Rate of NCC = −5%, Discharge Rate of MO = 10.5%). E. The peak infection time and patient distribution forecast of Hubei (Parameter Set: Decay Rate of NCC = −1.0%, Discharge Rate of MO = 17%). The peak infection time and patient distribution forecast of Hubei (Parameter Set: Decay Rate of NCC = −1.0%, Discharge Rate of MO = 10.5%).

## Discussion

In Hubei Province, since the outbreak of 2019-nCoV and the determination of human-to-human transmission, it has spread to all over China and even the world^10^. The growth trend of its cases is exponential during the epidemic period, but with the strict implementation of disease prevention and control, the inflection point of case growth will come as scheduled. Our results predicted the inflection point of patient growth and the distribution of patient status under different expectations by using mathematical models with Infectivity, Severity, and Lethality as indicators. The results show that under the existing medical and public health intervention, the Decay Rate of New Close Contacts is 10%, the inflection point of growth will come as early as February 23 (very optimistic), and the attenuation rate is 5% to 3 The beginning of the month (relatively optimistic), and when the decay rate drops to 1%, it will arrive at the latest on April 14 (very pessimistic).

As the birthplace of the disease, Hubei Province has mountains of confirmed and suspected cases. In order to finally overcome the disease, Hubei Province adopted plenty of even severe measures to restrict population movements, and adopted various strategies, such as prohibiting assembly, delaying school and resumption of work, or working from home, to significantly reduce the internal population contact rate. Although cases have spread throughout China, according to plans developed and implemented during previous major outbreaks such as SARS, the Middle East Respiratory Syndrome or a pandemic influenza outbreak, Hubei Province has eased intercity transmission through strict disease prevention and control and limits the worst cases to Hubei Province^11-13^. Persistent and compulsory implementation, as well as the cooperation of the people and the efforts of medical staff, will bring real relief. Through the guidance of the inflection point of the epidemic and the distribution of patient status, the government, medical work and tight medical resource allocation will become more efficient and reasonable. Wu, J. T. et al. indicated that it will peak in April with the infection rate staying put; it will reach a peak in May with the infection rate decreasing by 25%,; if the infection rate decreases to 50%, the epidemic will slowly increase in the first half of 2020 and will not reach its peak^9^.

The difficulty of the current epidemic control is that when these patients were admitted to the hospital, only 43.1% of them were feverish at the time, and more patients had fever during their hospitalization. A recent study showed that 2019-nCoV was detected in stool samples from patients with abdominal symptoms^14^. However, identifying and screening patients with atypical symptoms is difficult^15^. Rapid interpersonal transmission between close contacts is an important feature of 2019-nCoV pneumonia ^10, 16^. For SARS and Middle East Respiratory Syndrome (MERS), both of which are coronavirus-induced, almost all patients have fever symptoms when they are diagnosed, and only 1-2% does not have fever^17^. Thus, to promote the early inflection point, the infected people should be detected and isolated early, the medical team need to develop a new coronavirus vaccine, strengthened contact tracing and quarantine, and what’s more, wearing masks, avoiding crowd contact and strengthening their own immunity are requisite.

By utilizing the official notification data on the website of the National Health Commission, the State Transition Matrix Model predicted inflection points and patient distribution (based on the current state of the patient and not subject to historical factors). Owing to Hubei Province not affected by external factors such as people mobility (return peaks) and local control, compared to the national analysis, the effect only analyzing Hubei Province could be better. If the population is limited and the transition matrix is fixed, the above formulas will enough to predict all future results. However, the index of new cases will be affected by the input of PCR kit^18^. Simultaneously, as for confirming diagnosis of new cases, pharyngeal swabs are not as accurate as alveolar lavage fluid which means the possibility of false negatives. Furthermore, CT diagnosis is recommended, yet still with uncertainty^14^. Therefore, another advantage of this model is that, relative to the new cases, we choose to analyze critical and severe cases, which possess better stability, and the main medical resource consumption group is this group of patients. With the help of our model would optimize the critical medical resources allocation.

At present, tremendous medical resources are deployed in Wuhan, Hubei. If there is a slight slack in other areas, the prevention and control of the epidemic will be a devastating blow. Community governance is the basis of social governance and the most basic unit of national governance. In the comprehensive prevention and control system formed by the combination of national joint defense and joint control and group defense and group control, the community is an important coupling point, and the social epidemic prevention and protection system must be woven in the community. Only victory from Hubei can lead China to victory, and victory from China will bring the global victory.

Nevertheless, there are still limitations. Primarily, critical or severe cases will be affected by the gradually rational allocation of treatment teams and medical resources, the strengthening of medical power or virus toxicity decreasing. As a result, the peak time might arrive earlier than our estimates. Secondly, the impact of seasonal factors has not been considered and supposing 2019-nCoV is affected by the season, our prediction results may be unreliable. Hence, if these indicators continue to change, the model could have opportunities for improvement. In addition, due to the characteristics of severe and critical cases, there may be a huge impact on factors such as age and complex underlying diseases. Therefore, with the increasing awareness of the severity of the disease and the strengthening of prevention and control in various sectors of the society, various indicators may be reduced. At the same time, because of the instability of the previous indicators, these dynamic changes may affect the predictive efficacy of the model.

## Conclusions

We can infer that we are still not close to the end of this outbreak and the number of critically ill patients is still climbing. And assisting critical care resources in Hubei province requires the government to consider further tilt, and it is vital to make reasonable management of doctors and medical assistance systems to curb the transmission trend

## Data Availability

The availability of all data referred to in the manuscript

## Acknowledgement

Not applicable.

## Author’s contribution

Junhua Zheng and Chen Jian participated in study design; Ke Wu performed data analysis and drafted the manuscript; all author provided critical review of the manuscript and approved the final the final draft for publication.

## References

1. Chang, Lin M, Wei L, et al. Epidemiologic and Clinical Characteristics of Novel Coronavirus Infections Involving 13 Patients Outside Wuhan, China. JAMA. 2020.

2. Zhu N, Zhang D, Wang W, et al. A Novel Coronavirus from Patients with Pneumonia in China, 2019. N Engl J Med. 2020.

3. Heymann DL. Data sharing and outbreaks: best practice exemplified. Lancet. 2020.

4. Albright K, Gechter K, Kempe A. Importance of mixed methods in pragmatic trials and dissemination and implementation research. Acad Pediatr. 2013;13(5):400–7.

5. Coleman K, Austin BT, Brach C, Wagner EH. Evidence on the Chronic Care Model in the new millennium. Health Aff (Millwood). 2009;28(1):75–85.

6. Vijgen L, Keyaerts E, Moës E, et al. Complete genomic sequence of human coronavirus OC43: molecular clock analysis suggests a relatively recent zoonotic coronavirus transmission event. J Virol. 2005;79(3):1595–604.

7. de Wit E, van Doremalen N, Falzarano D, Munster VJ. SARS and MERS: recent insights into emerging coronaviruses. Nat Rev Microbiol. 2016;14(8):523–34.

8. Wang D, Hu B, Hu C, et al. Clinical Characteristics of 138 Hospitalized Patients With 2019 Novel Coronavirus-Infected Pneumonia in Wuhan, China. JAMA. 2020.

9. Wu JT, Leung K, Leung GM. Nowcasting and forecasting the potential domestic and international spread of the 2019-nCoV outbreak originating in Wuhan, China: a modelling study. Lancet. 2020.

10. Chan JF, Yuan S, Kok KH, et al. A familial cluster of pneumonia associated with the 2019 novel coronavirus indicating person-to-person transmission: a study of a family cluster. Lancet. 2020.

11. Ho PL, Tang XP, Seto WH. SARS: hospital infection control and admission strategies. Respirology. 2003;8 Suppl:S41–5.

12. Svoboda T, Henry B, Shulman L, et al. Public health measures to control the spread of the severe acute respiratory syndrome during the outbreak in Toronto. N Engl J Med. 2004;350(23):2352–61.

13. Cotten M, Watson SJ, Kellam P, et al. Transmission and evolution of the Middle East respiratory syndrome coronavirus in Saudi Arabia: a descriptive genomic study. Lancet. 2013;382(9909):1993–2002.

14. Song F, Shi N, Shan F, et al. Emerging Coronavirus 2019-nCoV Pneumonia. Radiology. 2020:200274.

15. Liu SL, Saif L. Emerging Viruses without Borders: The Wuhan Coronavirus. Viruses. 2020;12(2).

16. Phan LT, Nguyen TV, Luong QC, et al. Importation and Human-to-Human Transmission of a Novel Coronavirus in Vietnam. N Engl J Med. 2020.

17. Zumla A, Hui DS, Perlman S. Middle East respiratory syndrome. Lancet. 2015;386(9997):995–1007.

18. Corman VM, Landt O, Kaiser M, et al. Detection of 2019 novel coronavirus (2019-nCoV) by real-time RT-PCR. Euro Surveill. 2020;25(3).

